# Genome-wide association study stratified by *MAPT* haplotypes identifies potential novel loci in Parkinson’s disease

**DOI:** 10.1101/2023.04.14.23288478

**Authors:** Konstantin Senkevich, Sara Bandres-Ciga, Alejandro Cisterna-García, Eric Yu, Bernabe I. Bustos, Lynne Krohn, Steven J. Lubbe, Juan A. Botía, the International Parkinson’s Disease Genomics Consortium (IPDGC), Ziv Gan-Or

## Abstract

**Objective:** To identify genetic factors that may modify the effects of the *MAPT* locus in Parkinson’s disease (PD).

**Methods:** We used data from the International Parkinson’s Disease Genomics Consortium (IPDGC) and the UK biobank (UKBB). We stratified the IPDGC cohort for carriers of the H1/H1 genotype (PD patients n=8,492 and controls n=6,765) and carriers of the H2 haplotype (with either H1/H2 or H2/H2 genotypes, patients n=4,779 and controls n=4,849) to perform genome-wide association studies (GWASs). Then, we performed replication analyses in the UKBB data. To study the association of rare variants in the new nominated genes, we performed burden analyses in two cohorts (Accelerating Medicines Partnership – Parkinson Disease and UKBB) with a total sample size PD patients n=2,943 and controls n=18,486.

**Results:** We identified a novel locus associated with PD among *MAPT* H1/H1 carriers near *EMP1* (rs56312722, OR=0.88, 95%CI= 0.84-0.92, p= 1.80E-08), and a novel locus associated with PD among *MAPT* H2 carriers near *VANGL1* (rs11590278, OR=1.69 95%CI=1.40-2.03, p= 2.72E-08). Similar analysis of the UKBB data did not replicate these results and rs11590278 near *VANGL1* did have similar effect size and direction in carriers of H2 haplotype, albeit not statistically significant (OR= 1.32, 95%CI= 0.94-1.86, p=0.17). Rare *EMP1* variants with high CADD scores were associated with PD in the *MAPT* H2 stratified analysis (p=9.46E-05), mainly driven by the p.V11G variant.

**Interpretation:** We identified several loci potentially associated with PD stratified by *MAPT* haplotype and larger replication studies are required to confirm these associations.

## Introduction

Parkinson’s disease (PD) is a neurodegenerative disorder with complex genetic background, neuropathology, and clinical features. It is possible that what we define today as PD, which is based on clinical presentation only, is a combination of multiple disorders with different etiologies and pathologies but with similar clinical phenotype.^1^ PD is often categorized as an alpha-synucleinopathy, due to the typical accumulation of alpha-synuclein in Lewy bodies and Lewy neuritis.^2^ However, other pathologies such as tau pathology are also common in PD, as ∼50% of PD patients also show tau accumulation and deposition.^3^ In specific genetic subtypes of PD, such as *LRRK2*-associated PD, alpha-synuclein pathology can be found in only ∼50% of the cases,^4^ whereas tau pathology can be abundant in more than 90% of patients.^5^ In six brains from PD patients with the *LRRK2* p.I2020T mutation, only one had alpha-synucleinopathy, while tau pathology was very common.^6^ In contrast to *LRRK2*, in *GBA1*-associated PD, alpha-synuclein accumulation is very common, found in nearly all patients with *GBA1* variants, and tau pathology is likely less common.^7^ These genetic-neuropathological differences may suggest that tau is associated with specific genetic subtypes/variants.

Tau is encoded by *MAPT* (microtubule associated protein tau), a gene associated with multiple neurological disorders, including PD.^8^ The association with PD is mediated by the H1 and H2 *MAPT* haplotypes, created by an inversion of a large genomic region around *MAPT*.^9^ The H1 haplotype is the most common haplotype and H2 is more rare and absent in non-Caucasian populations.^10^ In PD, the H1 haplotype is associated with an increased risk of PD, whereas the H2 haplotype is associated with a reduced PD risk.^8^ The H1 haplotype and specific subhaplotypes have been linked to more severe tau pathology or increased tau expression in some neurodegenerative diseases,^11-13^ but whether or not these haplotypes affect tau pathology in PD is unclear. Recently, *MAPT* stratified analysis was done for Alzheimer’s disease,^14^ demonstrating potential novel loci and interactions.

Since *MAPT* encodes tau, which is found in brains of many PD patients, but with differential distribution in different genetic subtypes (e.g. *LRRK2* and *GBA1*), and since *MAPT* H1 and H2 haplotypes are associated with PD, we hypothesized that some genetic variants may be either more or less important in carriers of these haplotypes. To test this hypothesis, we performed stratified genome-wide association studies (GWAS) separately analyzing carriers of the H1/H1 genotype (PD patients n=8,492 and controls n=6,765) and carriers of the H2 haplotype (PD patients n=4,779 and controls n=4,849). We then examined whether specific variants are associated with PD in one of the genetically defined groups, but not in the other. To study the association of rare variants in the new nominated genes, we performed burden analyses in two cohorts with a total sample size PD patients n=2,943 and controls n=18,486.

## Methods

### Population and stratification

Our study included a total of 13,271 PD patients and 11,614 controls of European ancestry from the International Parkinson’s Disease Genomics Consortium (IPDGC) with available sex, age and principal components data. Details on the cohorts composing the IPDGC data can be found in Supplementary Table 1. To stratify by *MAPT* haplotype, we used the H2-tagging single nucleotide polymorphism (SNP) rs8070723 as previously reported. Carriers of the SNP rs8070723 are carriers of the H2 haplotype (with a genotype of either H1/H2 or H2/H2, PD patients n=4,779 and controls n=4,849), and non-carriers of this variant are carriers of the H1/H1 genotype (PD patients n=8,442 and controls n=6,765). The SNP rs8070723 is in high linkage disequilibrium (D’=1, r^2^=0.65) with the top SNP in the PD GWAS in the *MAPT* locus (rs62053943).

To study the role of rare variants in genes potentially interacting with *MAPT* from the previous analyses, we used whole genome and whole exome sequencing data from two cohorts: the Accelerating Medicines Partnership – Parkinson Disease (AMP-PD, 2.5 release; PD patients n=2,341 and controls n=3,486), and the UK biobank (UKBB; PD patients n=602 and controls n=15,000). The AMP-PD cohorts are detailed in the acknowledgments. We then stratified selected cohorts for rare variant analysis by *MAPT* haplotype, including H2 haplotype carriers (PD patients n=215, controls n=6,081 for UKBB and PD patients n=786, controls n=1,438 for AMP-PD) and H1/H1 haplotype carriers (PD patients n=387, controls n=8,919 for UKBB and PD patients n=1,555, controls n=2,048 for AMP-PD).

### Quality control and genome-wide association studies

Quality control procedures were performed for IPDGC genotyping data on individual and variant-level as previously described.^15^ Two case-control GWASs were performed using logistic regression in plink 1.9, including SNPs with a minor allele frequency (MAF) of > 0.01.^16^ We applied sex, age, and 10 principal components (PC) as covariates. One GWAS compared PD patients and controls who carry the *MAPT* H2 haplotype, and the second GWAS compared PD patients and controls who carry the H1/H1 genotype. We calculated genomic inflation factor (*λ*) for GWASs in both cohorts. To account for the impact of sample size, we scaled the *λ* values to 1000 cases and 1000 controls (*λ*_*1000*_) and created Quantile-Quantile plots as previously described.^17^ SNPs that surpass the Bonferroni threshold for genome-wide studies (5.0□×□10^−8^) were considered statistically significant. We created mirror Miami plots representing GWASs for carriers and non-carriers using the Hudson package in R (https://github.com/anastasia-lucas/hudson). Independent SNPs and loci were defined using the functional mapping and annotation portal (FUMA).^18^ To visually compare the effect size between carriers and non-carriers of the H2 haplotype, we constructed a beta-beta plot comparing the effect sizes in the two GWASs.

Variants with a difference of >5 fold in effect size were selected for further interaction analyses. We then examined whether interaction exists between the H2-tagging SNP rs8070723 and the SNPs that demonstrated differential effect sizes in the two GWASs. This analysis was done using plink and TLTO (Two-loci to OR) from VARI3 package.^19^

To replicate these potential interactions, we used the UK biobank data and performed stratification analysis followed by interaction analysis. Participants of European ancestry were selected from data field 22006. Heterozygosity outliers, samples with high missingness rates and samples with sex chromosome aneuploidy according to data field 22027/22019 were also excluded. The cohort also underwent filtration for relatedness using Genome-wide Complex Trait Analysis (GCTA) with a threshold of 0.125.^20^ In total, we included 344,597 unrelated participants of European origin, carriers of the H2 haplotype (PD patients n=550 and controls n=138,292), and carriers of the H1/H1 genotype (PD patients n=1,004 and controls n=204,749). The power to replicate results in UKBB was calculated using Genetic Association Study (GAS) power calculator.^21^ Locus zoom plots for regions of interest have been made using the online tool http://locuszoom.org/.^22^

### Rare variants burden analysis

We then studied rare variants in the novel genes that were nominated in the *MAPT* stratified GWASs. We performed quality control for whole-genome and whole-exome sequencing data using standard procedures on individual and variant levels as previously described.^23, 24^ Annotation was performed using ANNOVAR^25^ with Combined Annotation Dependent Depletion (CADD) score reference,^26^ where variants with a CADD score *≥*20 represent the top 1% of potentially deleterious variants. In this analysis, we only included variants with MAF <0.01. To perform burden analysis, we used an optimized sequence Kernel association test (SKAT-O and metaSKAT, R package).^27, 28^ Separate analyses were done for nonsynonymous variants and variants with high CADD score with adjustment for sex and age.

## Results

### Identification of variants associated with carriers and non-carriers of the *MAPT* H2 haplotype

To examine whether specific genetic variants are associated with carriers or non-carriers of the *MAPT* H2 haplotype, we performed two stratified GWASs. One GWAS included only H1/H1 genotype carriers (PD patients n=8,492 and controls n=6,765) and the second GWAS included carriers of either H1/H2 or H2/H2 genotypes, (PD patients n=4,779 and controls n=4,849). The genomic inflation values for H1 haplotype GWAS and H2/H2 haplotype GWAS were *λ* = 1.17 and *λ* = 1.11, respectively. When the genomic inflation was scaled to 1,000 cases and 1,000 controls for both cohorts, the values were the same, with *λ*_*1000*_=1.02 (Supplementary Figure 1). Overall, we identified seven loci in the *MAPT* H1/H1-stratified GWAS and six loci in the *MAPT* H2-stratified GWAS (Table 1, Figure 1A). In the *MAPT* H2-stratified GWAS, on top of known PD GWAS loci, we identified a novel locus associated with PD among carriers, tagged by rs11590278 (near *VANGL1*, OR=1.69, 95%CI=1.40-2.03, p=2.72E-08; Figure 1B). This SNP was not associated with PD in the H1/H1 stratified analysis (OR=1.06, 95%CI= 0.92-1.23, p=0.40). In the *MAPT* H1/H1-stratified GWAS we also identified a novel locus associated with PD in H1/H1 carriers, tagged by rs56312722 (near *EMP1*, OR=0.88, 95%CI= 0.84-0.92, p=1.80E-08; Figure 1C). This SNP was not associated with PD in the H2 stratified analysis (OR= 1.01, 95%CI= 0.96-1.07, p= 0.66). We also identified a novel locus in *SNAPC1*, which was significant in both carriers of H1/H1 haplotype rs113697492 (OR= 1.45, 95%CI= 1.29-1.63, p= 4.70E-10) and H2 haplotype rs75302411 (OR= 1.57, 95%CI= 1.37-1.80, p= 1.58E-10). Thus, this association was unrelated to haplotype subtyping. We did not find any effect of the novel loci from the *MAPT* stratified GWAS on age-at-onset of PD (Supplementary Table 2). Of the known PD loci, *GBA1, SNCA* and *TMEM175* were identified in both GWASs. The *NUCKS1* and *TMEM163* loci were associated with PD with the *MAPT* H1/H1 genotype, but not associated with PD with the H2 haplotype. Since the GWAS of the H1/H1 genotype is larger, this could be attributed to reduced power of the H2 haplotype GWAS, although the beta of the *NUCKS1* locus was more than two-fold larger in the H1/H1 genotype carriers compared to H2 haplotype carriers (−0.161 vs. -0.078, respectively). Similarly, the *MCCC1* locus was only associated with PD among carriers of the H2 haplotype, but not among carriers of the H1/H1. The beta for the SNP tagging the *MCCC1* locus, rs9858038, was more than two-fold larger in carriers of the H2 haplotype compared to carriers of the H1/H1 genotype (−0.224 vs. -0.108, respectively).

**Table 1.**
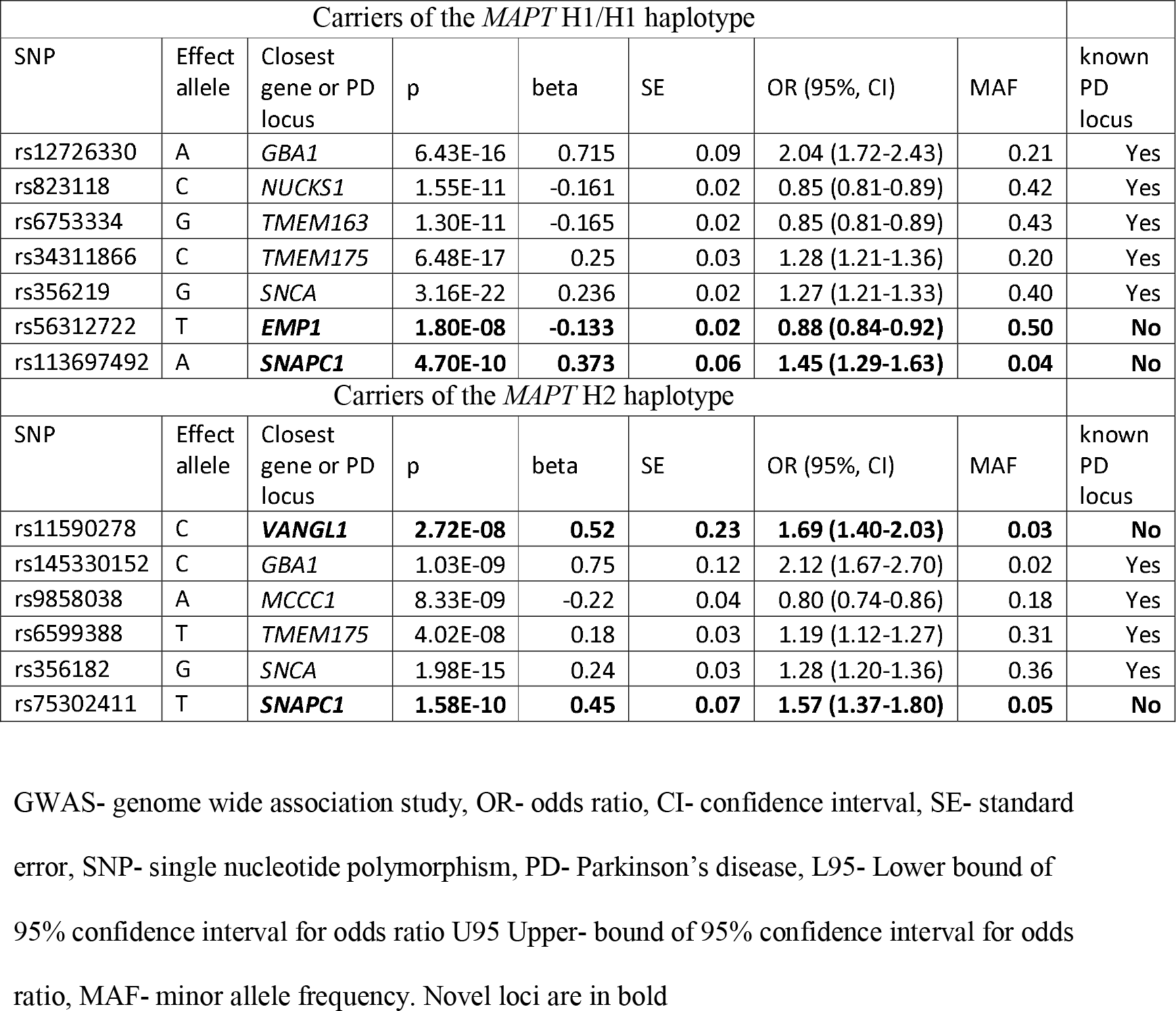
Significant loci in *MAPT* stratified GWAS.

| Carriers of the *MAPT* H1/H1 haplotype |  |  |  |  |  |  |  |  |
| --- | --- | --- | --- | --- | --- | --- | --- | --- |
| SNP | Effect allele | Closest gene or PD locus | p | beta | SE | OR (95%, CI) | MAF | known PD locus |
| rs12726330 | A | *GBA1* | 6.43E-16 | 0.715 | 0.09 | 2.04 (1.72-2.43) | 0.21 | Yes |
| rs823118 | C | *NUCKS1* | 1.55E-11 | -0.161 | 0.02 | 0.85 (0.81-0.89) | 0.42 | Yes |
| rs6753334 | G | *TMEM163* | 1.30E-11 | -0.165 | 0.02 | 0.85 (0.81-0.89) | 0.43 | Yes |
| rs34311866 | C | *TMEM175* | 6.48E-17 | 0.25 | 0.03 | 1.28 (1.21-1.36) | 0.20 | Yes |
| rs356219 | G | *SNCA* | 3.16E-22 | 0.236 | 0.02 | 1.27 (1.21-1.33) | 0.40 | Yes |
| rs56312722 | T | ***EMP1*** | **1.80E-08** | **-0.133** | **0.02** | **0.88 (0.84-0.92)** | **0.50** | **No** |
| rs113697492 | A | ***SNAPC1*** | **4.70E-10** | **0.373** | **0.06** | **1.45 (1.29-1.63)** | **0.04** | **No** |
| Carriers of the *MAPT* H2 haplotype |  |  |  |  |  |  |  |  |
| SNP | Effect allele | Closest gene or PD locus | p | beta | SE | OR (95%, CI) | MAF | known PD locus |
| rs11590278 | C | ***VANGL1*** | **2.72E-08** | **0.52** | **0.23** | **1.69 (1.40-2.03)** | **0.03** | **No** |
| rs145330152 | C | *GBA1* | 1.03E-09 | 0.75 | 0.12 | 2.12 (1.67-2.70) | 0.02 | Yes |
| rs9858038 | A | *MCCC1* | 8.33E-09 | -0.22 | 0.04 | 0.80 (0.74-0.86) | 0.18 | Yes |
| rs6599388 | T | *TMEM175* | 4.02E-08 | 0.18 | 0.03 | 1.19 (1.12-1.27) | 0.31 | Yes |
| rs356182 | G | *SNCA* | 1.98E-15 | 0.24 | 0.03 | 1.28 (1.20-1.36) | 0.36 | Yes |
| rs75302411 | T | ***SNAPC1*** | **1.58E-10** | **0.45** | **0.07** | **1.57 (1.37-1.80)** | **0.05** | **No** |

GWAS- genome wide association study, OR- odds ratio, CI- confidence interval, SE- standard error, SNP- single nucleotide polymorphism, PD- Parkinson's disease, L95- Lower bound of 95% confidence interval for odds ratio U95 Upper- bound of 95% confidence interval for odds ratio, MAF- minor allele frequency. Novel loci are in bold

**Figure 1.**
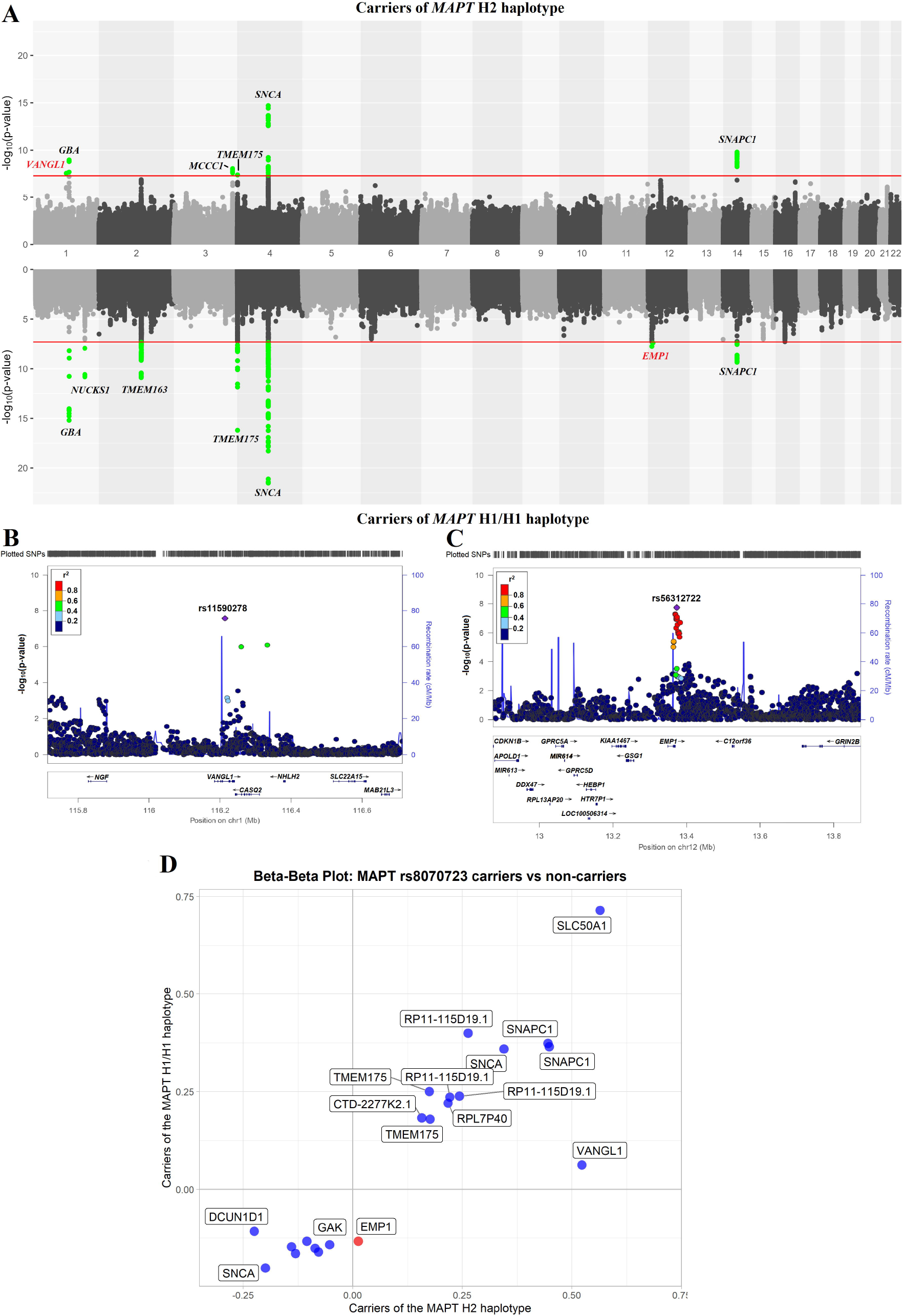
**A.** Miami plot of carriers (top) and non-carriers (bottom) of *MAPT* H2 haplotype of selected variants. The red line indicates GWAS significance threshold. Green dots indicate passing the threshold variants. New loci with potential interaction with *MAPT* highlighted in red, other loci highlighted in black. **B.** Locus zoom plot of *VANGL1* variants (+/- 500kb) in the stratified PD risk GWAS of the *MAPT* H2 haplotype carriers. **C.** Locus zoom plot of *EMP1* variants (+/- 500kb) in the stratified PD risk GWAS of the MAPT H1/H1 haplotype carriers. The position of the variants on the chromosome (x axis) is plotted against the log10-scaled p-values (left y axis). SNP with the smallest p-value is indicated by a purple diamond. Linkage disequilibrium scores related to the top variant are defined by different colors, explained by the legend on the upper left corner. The right vertical axis indicates the regional recombination rate (cM/Mb). Abbreviations: chr, chromosome; cM, centimorgan; Mb, Megabase; PD, Parkinson’s disease; SNP, single nucleotide polymorphism. **D.** Beta-beta plots for independent SNPs that reached genome wide association level of significance in carriers or non-carriers of *MAPT* rs8070723 (Carriers of the *MAPT* H2 haplotype vs carriers of the *MAPT* H1/H1 haplotype). Red circles denote SNPs with effect size (beta) difference of more than 5 times between carriers of *MAPT* H2 haplotype and carriers of the *MAPT* H1/H1 haplotype.

We attempted to replicate these results, specifically the novel loci, in the UKBB data, stratified by carriers and non-carriers of the *MAPT* H2 haplotype. Considering the number of *MAPT* haplotype carriers, allele frequency and effect size of novel loci, we calculated the statistical power for replication. We did not have sufficient power to replicate neither rs11590278 (60%) nor rs56312722 (54%) and as expected both associations did not reach statistical significance in the UKBB. However, the SNP rs11590278 near *VANGL1* had similar effect direction in carriers of H2 haplotype (OR= 1.32, 95%CI= 0.94-1.86, p=0.17). The SNP rs56312722 near *EMP1* had different direction of effect in carriers of H1/H1 haplotype (OR= 1.07, 95%CI= 0.97-1.16, p= 0.10).

### Interaction analysis of *MAPT* H1 and H2 haplotypes with SNPs in the *VANGL1* and *EMP1* loci

To identify SNPs for interaction analyses with the *MAPT* H2-tagging SNP rs8070723, we generated a beta-beta plot (Figure 1D). We selected independent SNPs that had an effect size (beta) >5-fold larger in one GWAS compared to the other and were significant in the GWAS level in one of the two GWASs. The SNP rs11590278 (*VANGL1*) had an 8-fold difference in effect size when comparing *MAPT* H2 haplotype carriers and H1/H1 genotype carriers (b=0.52, p=2.72E-08 vs b=0.062, p=0.4, respectively). The SNP rs56312722 *(EMP1*) had a ten-fold difference in beta compared to H2 haplotype carriers (b=0.133, p=1.80E-08 vs b=0.013, p=0.6582, respectively).

We then performed allele-specific interaction in a stratified cohort between *MAPT* H2-tagging SNP rs8070723 and the SNPs of interest (rs11590278 *VANGL1* and rs56312722 *EMP1;* Table 2). We found that C/T allele of rs11590278 (*VANGL1*) interacted with H1/H2 *MAPT* haplotype (OR=1.73, 95%CI=1.40-2.18, p=9.93E-08). The direction of the effect was preserved in the UKBB data but without statistical significance (OR=1.40, 95%CI=0.94-2.01, p=0.083). In the subsequent analysis, we found an interaction between rs56312722 *EMP1* T/T genotype and *MAPT* H1/H1 haplotype (OR=0.84, 95%CI=0.78-0.91, p=5.34E-06). This interaction was not replicated in UKBB (OR=1.12, 95%CI=0.97-1.30, p=0.101).

**Table 2.**
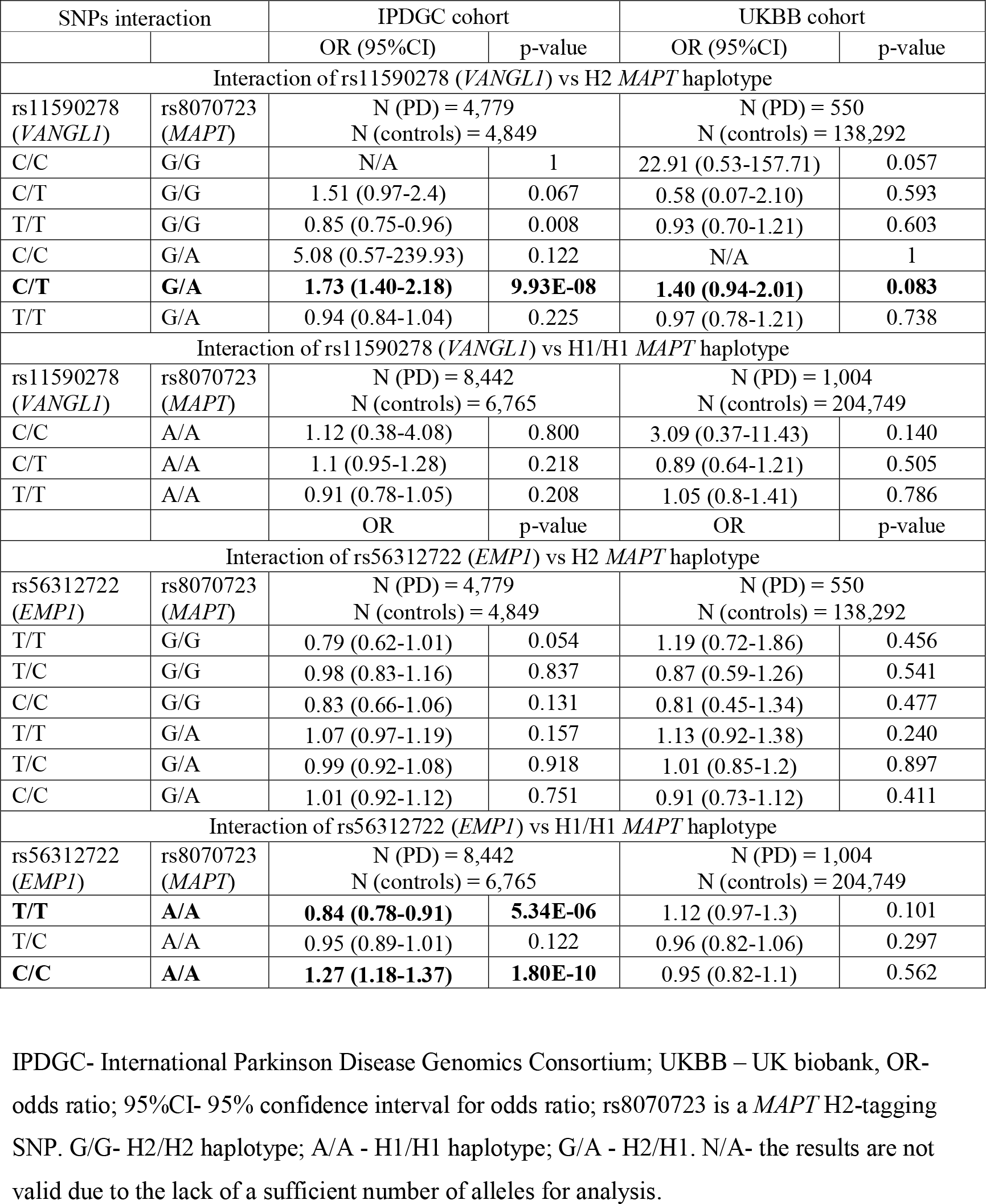
Allele specific interaction between *MAPT* H2-tagging SNP rs8070723 two new loci (rs11590278 *VANGL1* and rs56312722 *EMP1*)

### Rare variant analysis further suggests an association of *EMP1* with Parkinson’s disease

To examine whether rare variants in *EMP1* and *VANGL1* play any role in PD, we performed burden analysis of variants with MAF<0.01. We found that rare variants with CADD score >20 were associated with PD in the meta-analysis of AMP-PD and UKBB cohorts (p=0.004; Supplementary Table 3). This association was driven by the UKBB cohort (p=0.0004). Moreover, after we stratified the cohorts for H2 and H1/H1 *MAPT* haplotypes the same way we did for the GWAS, we found that the association of *EMP1* with PD was driven by the H2 haplotype carriers in both AMP-PD (p=0.018) and UKBB (p=0.004) cohorts (meta-analysis p=9.46E-05). There were only two variants with high CADD score and only the p.V11G *EMP1* variant was associated with PD in the UKBB cohort (OR=8.62, 95%CI=2.21-33.61; p=0.002) and in AMP-PD (OR=5.92, 95%CI=1.12-31.14, p=0.036) in the H2 stratified cohort (Supplementary Table 4). Furthermore, p.V11G was in linkage disequilibrium with the major allele of rs56312722 in *EMP1* (D’=1.0). We did not find any association between rare variants in *VANGL1* and PD in any of the cohorts.

## Discussion

In the current study we performed a stratified GWAS based on the *MAPT* H1 and H2 haplotypes, and identified three novel loci potentially associated with PD, one in carriers of the H1/H1 genotype (*EMP1*), one in carriers of the H2 haplotype (*VANGL1*), and one in both (*SNAPC1*). These associations were not replicated in the UKBB data, which was underpowered to detect them. We also found that rare *EMP1* variants were associated with PD in H2 haplotype carriers in both cohorts (AMP-PD and UKBB). Therefore, possibly there is a complex interaction between *MAPT* haplotypes and the *EMP1* gene, with some variants being protective for H1/H1 haplotype and causative for H2 haplotype. Additional studies are needed to examine whether the *VANGL1* and *EMP1* loci are indeed associated with the H2 and H1 haplotypes in PD. As for the third locus, *SNAPC1*, it was not identified in a much larger GWAS,^15^ suggesting that this could be a spurious association. However, it is also possible that methodological and/or population stratification-related issues due to inclusion of data from 23andMe masked the association in the larger GWAS. More studies are therefore needed into this locus to determine whether or not it is associated with PD.

*MAPT* stratified analysis has already been performed in Alzheimer’s disease,^14^ and an *APOE*-stratified GWAS has been recently performed in dementia with Lewy bodies,^29^ both identified potential novel loci and interactions. Together with our current work, these studies demonstrate the advantage of a stratified GWAS approach for shedding more light on the genetics of complex neurological traits with multiple potential subtypes.

In the stratified analyses by *MAPT* H1 and H2 haplotypes, we nominated two novel loci potentially associated with PD, near the *VANGL1* and *EMP1* genes. *EMP1* is encoding epithelial membrane protein 1 and this gene has not previously been implicated in the pathogenesis of PD. In a recent RNA-seq study of cerebral white matter, upregulation of *EMP1* was reported in multiple system atrophy patients.^30^ *GRIN2B*, located in the *EMP1* locus, was previously associated with impulse control and related behaviors in PD patients.^31^ The *MAPT* H1 haplotype has been repeatedly associated with cognitive decline in PD patients.^32-34^ Likewise, the *EMP1* locus has been previously associated with 176 traits including intelligence and cognitive performance (data retrieved from https://genetics.opentargets.org/). Thus, *EMP1* could be an independent modifier of PD in *MAPT* H1 or H2 carriers. However, this hypothesis requires additional studies.

*VANGL1* encodes the Van Gogh-like planar cell polarity protein 1 and was previously associated with neural tube defects,^35, 36^ but not with neurodegenerative diseases. *VANGL1* is a negative regulator of the canonical Wnt signaling pathway.^37^ Notably, Wnt signaling has been suggested to be an important factor for dopaminergic neurons homeostasis and survival.^38, 39^ Therefore, the involvement of *VANGL1* in PD pathogenesis should also be further studied. Both genes, *EMP1* and *VANGL1*, are expressed in brain tissues based on GTEx (data retrieved from https://www.gtexportal.org/home). However, we did not find quantitative trait loci for SNPs in *EMP1* and *VANGL1* associated with PD in the current study.

The *KANSL1* gene has recently been nominated as potentially driving an association for the H1 haplotype at the *MAPT* locus.^40^ Additionally, it has been suggested that *KANSL1* and *KAT8* are two PINK1-dependent mitophagy regulators.^41^ In our analysis, *KAT8* almost reached genome-wide association significance in the H1/H1 haplotype analysis (p=5.21E-08) and was only nominally significant in the H2 haplotype analysis (p=0.0004), with no difference in effect direction (Figure 1A). It is possible that this is due to an interaction between *KAT8* and the H1 haplotype, but it could also be a power issue as the GWAS with H1/H1 carriers is much larger than H2. Therefore, further analysis is needed to determine whether *KAT8* locus does indeed interact with the *MAPT* H1 haplotype.

Our analyses have several limitations. First, we have separated the discovery IPDGC cohort into smaller groups of carriers and non-carriers which caused imbalance in group size depending on the variant frequency and, thus, different power. We have also performed replication analyses in UKBB, which only had 550 PD H2 haplotype carries and 1,004 PD H1/H1 haplotype carriers after filtering. This makes UKBB underpowered to detect the effects seen in our discovery cohorts. Since we only studied European samples, similar studies in non-European cohorts are warranted, as well as replication of our findings in independent and larger cohorts. Another limitation is that we had several GWAS-significant loci with low MAF < 5%. Therefore, these signals require cautious interpretation and further confirmation.

To conclude, using a stratified GWAS approach, we nominated two potentially novel loci which are associated with specific *MAPT* gene haplotypes in PD. Our results along with previous similar studies suggest that risk loci stratification analysis could help us to genetically characterize subtypes of PD patients better and discover novel genetic associations.

## Data Availability

All data produced in the present study are available upon reasonable request to the authors and the code used in the study is available on github

https://github.com/gan-orlab/PD_stratified_GWAS

## Data Availability

Full code used in the analysis has been made available on git-hub (https://github.com/gan-orlab/PD_stratified_GWAS).

## Acknowledgements

We would like to thank the all the participants in the different cohorts. We would also like to thank all members of the International Parkinson Disease Genomics Consortium (IPDGC). See for a complete overview of members, acknowledgements, and funding http://pdgenetics.org/partners. This research has been conducted using the UK Biobank Resource under Application Number 45551. This research used the NeuroHub infrastructure and was undertaken thanks in part to funding from the Canada First Research Excellence Fund, awarded through the Healthy Brains, Healthy Lives initiative at McGill University, Calcul Québec and Compute Canada. Data used in the preparation of this article were obtained from the AMP PD Knowledge Platform. For up-to-date information on the study, visit https://www.amp-pd.org. AMP PD – a public-private partnership – is managed by the FNIH and funded by Celgene, GSK, the Michael J. Fox Foundation for Parkinson’s Research, the National Institute of Neurological Disorders and Stroke, Pfizer, Sanofi, and Verily. Genetic data used in preparation of this article were obtained from the Fox Investigation for New Discovery of Biomarkers (BioFIND), the Harvard Biomarker Study (HBS), the Parkinson’s Progression Markers Initiative (PPMI), the Parkinson’s Disease Biomarkers Program (PDBP), the International LBD Genomics Consortium (iLBDGC), and the STEADY-PD III Investigators. BioFIND is sponsored by The Michael J. Fox Foundation for Parkinson’s Research (MJFF) with support from the National Institute for Neurological Disorders and Stroke (NINDS). The BioFIND Investigators have not participated in reviewing the data analysis or content of the manuscript. For up-to-date information on the study, visit michaeljfox.org/news/biofind. The HBS is a collaboration of HBS investigators [full list of HBS investigator found at https://www.bwhparkinsoncenter.org/biobank/ and funded through philanthropy and NIH and Non-NIH funding sources. The HBS Investigators have not participated in reviewing the data analysis or content of the manuscript. PPMI – a public-private partnership – is funded by the Michael J. Fox Foundation for Parkinson’s Research and funding partners, including [list the full names of all of the PPMI funding partners found at www.ppmi-info.org/fundingpartners]. The PPMI Investigators have not participated in reviewing the data analysis or content of the manuscript. For up-to-date information on the study, visit www.ppmi-info.org. PDBP consortium is supported by the NINDS at the National Institutes of Health. A full list of PDBP investigators can be found at https://pdbp.ninds.nih.gov/policy. The PDBP investigators have not participated in reviewing the data analysis or content of the manuscript. Genome Sequencing in Lewy Body Dementia and Neurologically Healthy Controls: A Resource for the Research Community.” was generated by the iLBDGC, under the co-directorship by Dr. Bryan J. Traynor and Dr. Sonja W. Scholz from the Intramural Research Program of the U.S. National Institutes of Health. The iLBDGC Investigators have not participated in reviewing the data analysis or content of the manuscript. For a complete list of contributors, please see: bioRxiv 2020.07.06.185066; doi: https://doi.org/10.1101/2020.07.06.185066. STEADY PD III is a 36 month, Phase 3, parallel group, placebo controlled study of the efficacy of isradipine 10 mg daily in 336 participants with early Parkinson’s Disease that was funded by the NINDS and supported by The Michael J Fox Foundation for Parkinson’s Research and the Parkinson’s Study Group. The STEADY-PD III Investigators have not participated in reviewing the data analysis or content of the manuscript. The full list of STEADY PD III investigators can be found at: https://clinicaltrials.gov/ct2/show/NCT02168842. ZGO is supported by the Fonds de recherche du Québec - Santé (FRQS) Chercheurs-boursiers award, and is a William Dawson Scholar. KS is supported by a postdoctoral fellowship from the Canada First Research Excellence Fund (CFREF), awarded to McGill University for the Healthy Brains for Healthy Lives initiative (HBHL) and postdoctoral fellowship from FRQS. AC was supported by the Science and Technology Agency, Séneca Foundation, Comunidad Autónoma Región de Murcia, Spain through the grant 20762/FPI/18. This work was supported in part by the Intramural Research Programs of the National Institute of Neurological Disorders and Stroke (NINDS) and the National Institute on Aging (NIA). This study was financially supported by grants from the Michael J. Fox Foundation, the Canadian Consortium on Neurodegeneration in Aging (CCNA), the Canada First Research Excellence Fund (CFREF), awarded to McGill University for the Healthy Brains for Healthy Lives initiative (HBHL), and Parkinson Canada.

## Author Contributions

Concept and design: KS, ZGO

Data acquisition: KS, SBS, ACG, EY, BIB, LK, SJL, JAB, ZGO

Statistical analysis: KS, SBS, ACG, ZGO

Drafting of the manuscript: KS, ZGO

Critical revision of the manuscript: All

## Financial Disclosure

Authors have nothing to report.

## Supplementary files

Supplementary Figure 1. Quantile-quantile (Q-Q) plot for MAPT stratified GWAS. A. Q-Q plot of MAPT H1/H1 haplotype carriers B. Q-Q plot of MAPT H2 haplotype carriers.

Supplementary Table 1. Demography of studied population.

Supplementary Table 2. Effect of novel SNPs EMP1 (rs56312722) and VANGL1 (rs11590278) variants on Parkinson’s disease age-at-onset in MAPT stratified cohorts.

Supplementary Table 3. Burden analysis of rare *EMP1* and *VANGL1* variants in MAPT stratified cohorts.

Supplementary Table 4. *EMP1* variants with high CADD score.

## Notes

### Competing Interest Statement

The authors have declared no competing interest.

### Funding Statement

FRQS, HBHL, CFREF, MJFF, Intramural Research Programs of the National Institute of Neurological Disorders and Stroke (NINDS) and the National Institute on Aging (NIA).

### Author Declarations

Ethics committee/IRB of McGill University gave ethical approval for this work

