## Supplementary figures and images for "Genome-wide association study stratified by *MAPT* haplotypes identifies potential novel loci in Parkinson’s disease"

### Supplementary Figure

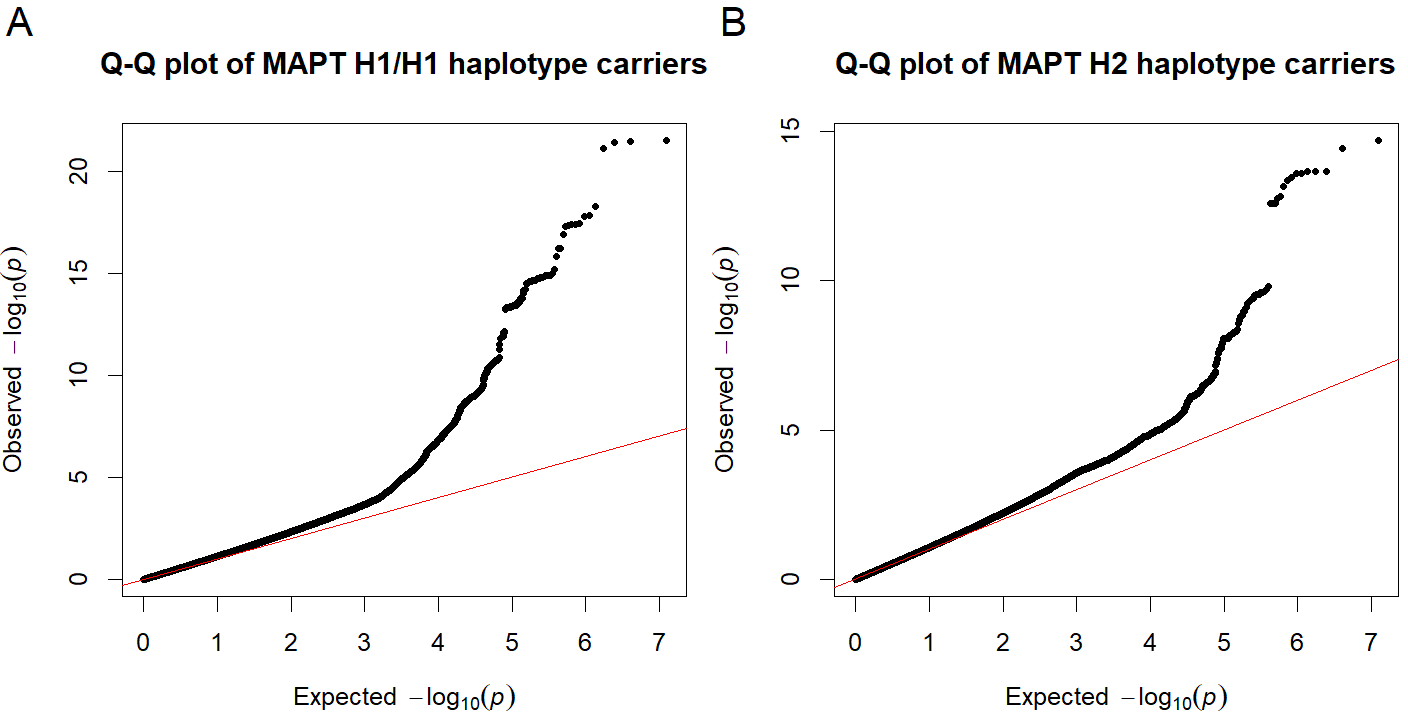
